# Optimizing the Implementation of Clinical Predictive Models to Minimize National Costs: A Sepsis Case Study

**DOI:** 10.1101/2022.08.28.22279313

**Authors:** Parker Rogers, Aaron E. Boussina, Supreeth P. Shashikumar, Gabriel Wardi, Christopher A. Longhurst, Shamim Nemati

**Affiliations:** Department of Economics, University of California San Diego, San Diego, USA; Department of Biomedical Informatics, University of California San Diego, San Diego, USA; Department of Emergency Medicine, University of California, San Diego, San Diego, California; Division of Pulmonary, Critical Care, and Sleep Medicine, University of California-San Diego, San Diego, California

**Keywords:** sepsis, machine learning, evaluation, utility assessment, workflow simulation

## Abstract

**Objective:** To optimize the parameters of a sepsis prediction model within distinct patient groups to minimize the excess cost of sepsis care and analyze the potential effect of factors contributing to end-user response to sepsis alerts on overall model utility.

**Materials and Methods:** We calculated the excess costs of sepsis by comparing patients with and without a secondary sepsis diagnosis but with the same primary diagnosis and baseline comorbidities. We optimized the parameters of a sepsis prediction algorithm across different diagnostic categories to minimize these excess costs. At the optima, we evaluated diagnostic odds ratios and analyzed the impact of compliance factors—like non-compliance, treatment efficacy, and tolerance for false alarms—on the net benefit of triggering sepsis alerts.

**Results:** Compliance factors significantly contributed to the net benefit of triggering a sepsis alert. However, a customized deployment policy can achieve a significantly higher diagnostic odds ratio and reduced costs of sepsis care. Implementing our optimization routine with powerful predictive models could result in $4.6 billion in excess cost savings for the Medicare program.

**Discussion:** Sepsis costs and incidence vary dramatically across diagnostic categories, warranting a customized approach for implementing predictive models. We designed a framework for customizing sepsis alert protocols within different diagnostic categories to minimize excess costs and analyzed model performance as a function of false alarm tolerance and compliance with model recommendations.

**Conclusion:** Customizing the implementation of clinical predictive models by accounting for various behavioral and economic factors may improve the practical benefit of predictive models.

## INTRODUCTION

Recent advancements in machine learning (ML) and the proliferation of healthcare data have led to widespread excitement about using these technologies to improve care^1,2^. Predictive analytic models in domains such as sepsis^3–5^, acute kidney injury^6^, respiratory failure^7^, and general deterioration^8^ have been proposed to improve the timely administration of life-saving treatments and mitigate expensive downstream complications. It has been argued that a more tailored approach that accounts for implementation constraints that may differ across care settings can further enhance the adoption of such systems^9^.

Despite its importance, the process of implementing predictive analytics solutions has received little attention relative to the development of the underlying ML models^10^. Algorithms are becoming more sophisticated, and the infrastructure that allows real-time, interoperable deployment of predictive analytics solutions is expanding^11,12^. This increase in potential and complexity underscores the practical importance of understanding the implementation policy layer, which captures the clinical workflow, response protocols, and operational constraints. Notably, the dominant evaluation methods within the ML community, such as the area under the receiver-operator curve (AUROC), often do not consider the effect of this policy layer on model performance^13^. Moreover, such performance metrics do not consider the user response to prediction and the effectiveness of the treatment protocols^14^. However, the operational constraints can often go beyond behavioral factors and may encompass quality improvement (QI) mandates and cost-saving objectives^15^.

This work focuses on the management of sepsis, a common and lethal condition caused by a dysregulated host response to infection^16^, though our framework can be applied to other hospital-acquired conditions^17^. Sepsis afflicts over 49 million people worldwide and accounts for over 11 million deaths per year^18^. In 2018, the U.S. Medicare program (including fee-for-service and Medicare Advantage) incurred $41.5 billion dollars in sepsis-related inpatient hospital admissions and skilled nursing facility care costs^19^.

We propose a framework for improving the implementation of ML-based EHR alerts. Our framework aims to minimize the costs of sepsis that are avoidable through early detection, timely administration of antibiotics, and prevention of overtreatment (i.e., “excess costs”) ^5,20,21^. Importantly, these costs can differ by diagnostic category due to differences in incidence rates, patient susceptibility, and physician adherence. Little is known, however, of the magnitude of these excess costs in inpatient settings. Thus, an additional contribution of this work is our estimation of the excess costs of sepsis at the diagnostic-category and national level (i.e., costs paid by the Medicare program). Our optimization framework uses these cost estimates and selects specific decision thresholds for each diagnostic category, differing from other cost-benefit frameworks that set decision thresholds uniformly^22,23^. This tailored approach results in higher cost savings and diagnostic accuracy.

## MATERIALS AND METHODS

We conducted a retrospective observational study with the following three broad steps: data collection, excess cost estimation, and cost minimization (Figure 1). This was done in accordance with STROBE guidelines^24^.

**Figure 1.**
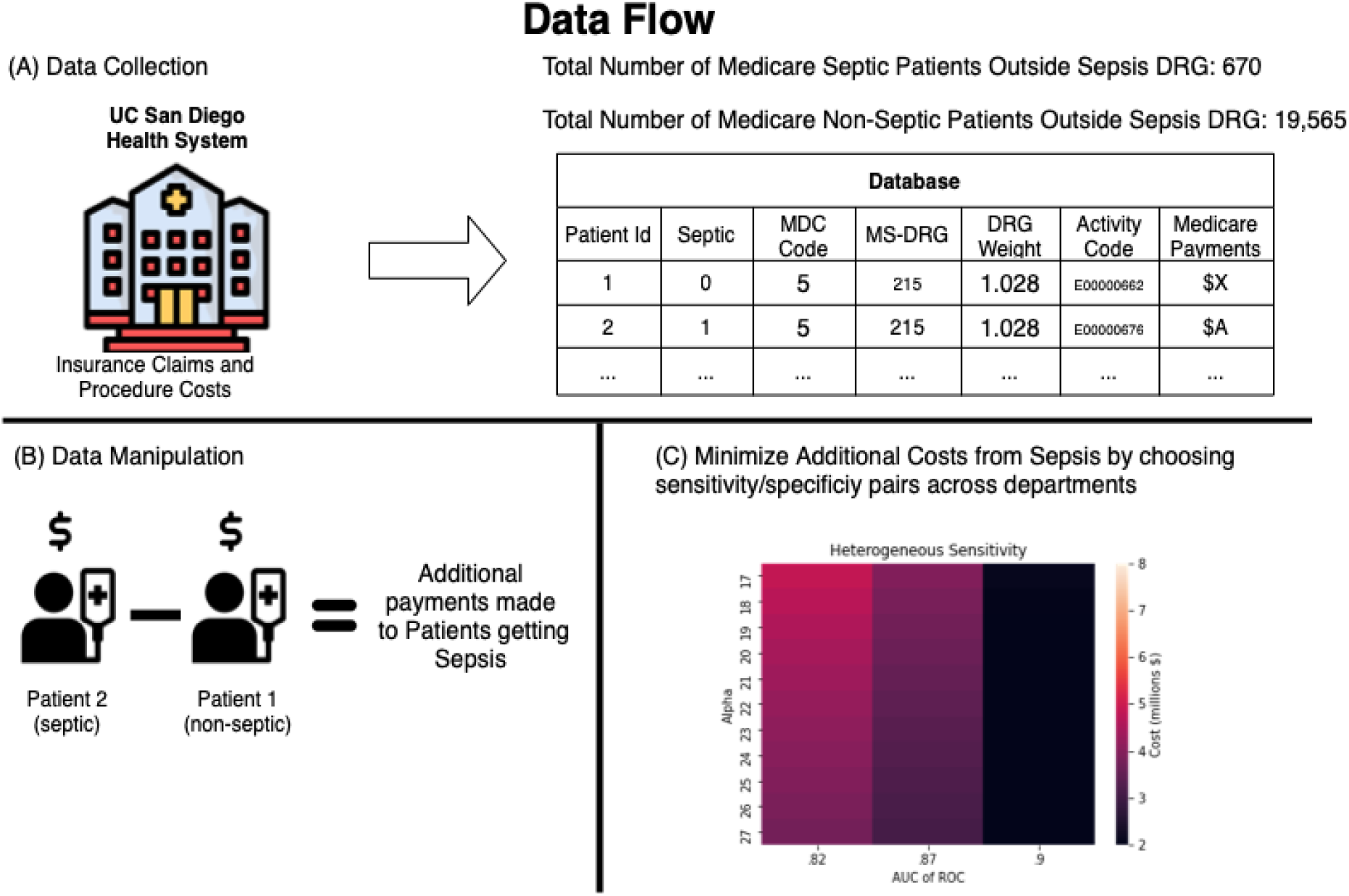
Overall framework for assessment of attributable cost to sepsis and optimization of predictive model parameters.

### Datasets and Definitions

The Institutional Review Board (IRB) of UC San Diego approved this study (#800257) with a waiver of informed consent. We collected insurance claims data from adult patients at UC San Diego Health, an academic health system, between October 2016 and July 2020. These data included the following necessary components: (i) the Medicare Severity Diagnosis Related Groups (MS-DRGs) diagnosis code for each patient and their corresponding DRG weights, (ii) the total amount paid by Medicare for the patient, and (iii) the Charlson Comorbidity Index (CCI) of the patient upon admission. We included patients with International Classification of Disease 10 (ICD 10) codes for severe sepsis (ICD9: 99592 and ICD10: R6520) and septic shock (ICD9: 78552 and ICD10: R6521). We selected these because of their inclusion in the Centers for Medicare and Medicaid Service (CMS) Quality Measure for Severe Sepsis and Septic Shock (SEP1), which has impacted sepsis care across the United States and provides a standardized approach to management^25^. Throughout the manuscript, the term “sepsis” refers to these definitions of severe sepsis and septic shock.

### Excess Cost of Sepsis

Our efforts to quantify the costs of missed diagnoses (i.e., false negatives) provide a new estimate of the avoidable costs of severe sepsis and septic shock across broad diagnostic categories. To quantify, we used granular insurance claims data under the Medicare prospective payment system (PPS). We focused on hospitalized Medicare patients as payments are specific to Diagnosis Related Groups (DRGs), a payment classification system that is determined primarily by the diagnosis that caused a patient to become hospitalized^26^. This system groups clinically similar conditions that require similar levels of inpatient resources. This categorization also allows us to show the public value of our optimization routine. We excluded patients from sepsis related DRGs (870, 871, 872) from our analysis since our objective is to assess the *excess inpatient cost* of sepsis for other DRGs. As such, we gathered all severe sepsis and septic shock patients in non-sepsis DRGs and a group of control patients in those same DRGs. This strategy allowed a cost comparison between individuals with similar primary diagnoses (i.e., underlying conditions) but different secondary sepsis diagnoses. These data included 670 patients diagnosed with severe sepsis and septic shock across 131 DRGs and 19,565 control group patients.

We adjusted for other underlying factors that drive cost differences between septic and non-septic patients by matching a comparison individual to each septic patient^27^. For each septic patient, this matching procedure selected from all comparison individuals within the same DRG code/weight the patient with the most similar CCI to the given septic patient^1^. Further, limiting the selection to patients within the same DRG weight accounts for changes in DRG payments over time. With sets of septic patients matched to control patients, we differenced the Medicare payments for the septic patients and the matched patients. This difference represents the excess costs that Medicare paid for sepsis above the underlying costs attributable to the primary patient diagnosis.

We repeat this procedure for all septic patients to form a distribution of excess costs across DRGs. We then average these DRG-specific excess cost estimates by Major Diagnostic Categories (MDC), which are comprised of 16 mutually exclusive diagnosis areas within our dataset^28^. We show that the added costs from sepsis diagnoses vary dramatically by these diagnostic categories and, by extension, different hospital departments.

In effort to calculate the national excess cost of sepsis, we then scale our excess cost estimates to the national level to show the public impact of early detection and treatment (see Supplementary Material for more details). To scale, we first multiply UC San Diego Health (UCSDH) payments by the ratio of UCSDH payments to average U.S. payments by DRG. Then we scale the total patients treated at UCSDH to the national number of septic patients using the share of Medicare patients treated at UCSDH. Lastly, we aggregate the payments across all patients. We validate this scaling approach in the Supplementary Materials section and find that we closely estimate the total national inpatient sepsis costs documented in Medicare cost data^29^ (among sepsis DRGs 870-872) by scaling UCSDH total sepsis costs to the national level.

### Modeling and Optimization

We use the sepsis prediction model by Shashikumar at al.^5^ to develop an optimization framework that chooses the model’s classification thresholds to minimize the additional costs from sepsis by the MDCs. In the context of sepsis prediction, classification thresholds determine above which probability the model tags a patient as septic. Although we optimize across diagnostic categories, our routine could also be implemented across hospital departments or, alternatively, across more granular patient subpopulations. The intuition behind the value of our implementation rests on the idea that septic patients may be more (less) costly across diagnostic categories, which could potentially merit a lower (higher) classification threshold. Additionally, departments could have different rates of sepsis which may require different thresholds to avoid a large number of missed detections. By allowing algorithmic sensitivity to adjust to these idiosyncrasies, ML algorithms may further reduce costs. Our optimizer is constrained by the predictive model’s AUC: as the optimizer chooses a higher sensitivity to sepsis to reduce the costs of sepsis, the specificity of the model decreases, increasing the false alarm rate. As noted above, false alarms can also be costly: treating patients with broad-spectrum antibiotics can cause adverse effects and is expensive. Thus, the algorithm must balance the trade-off between the cost of under-treatment and over-treatment.

Let *FN*_*i*_ represent the number of false negatives (i.e., the missed cases of sepsis) within a given MDC category, and Cost_*FN*_*i*_ represent the cost of missing sepsis within this category. The miss rate (i.e., *1 – sens*_*i*_) is a quantity that depends on the selection of risk score threshold within the given MDC category *i*. Furthermore, let the estimated functional form *f()* provide a mapping from the chosen sensitivity to the false positive rate (FPR; or false alarm rate). Note that this function is constrained by the model AUC and reflects the balance of model sensitivity and false alarms (see Supplementary Material, section 5).

The optimization routine is given by:

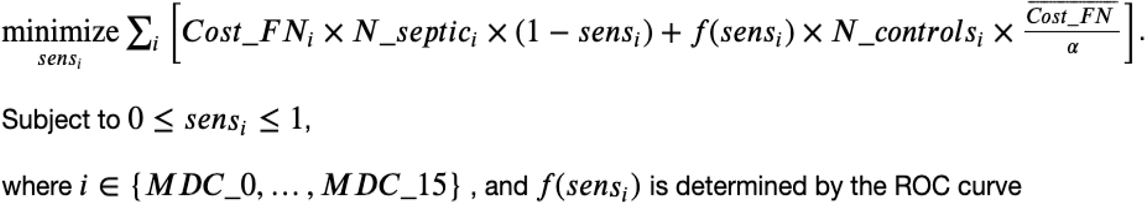

Notice that the algorithm chooses sensitivity values (i.e., *sens*_*i*_) across 16 broad diagnostic categories (i.e., *i*) to minimize costs. The left-hand side of the objective function captures the excess costs, or the cost of false negatives: the MDC’s average cost of false negatives, multiplied by the number of septic patients in the diagnostic category (i.e., *N_septic*_*i*_), multiplied by the miss rate. The right-hand side of the objective function captures the costs of false positives: the false alarm rate, multiplied by the number of patients who are not septic in the MDC (i.e., *N_controls*_*i*_), multiplied by the average costs of false negatives divided by the conversion factor *α*. This conversion factor is a variable that maps the cost of false positives to the cost of false negatives as there may be costs of overtreatment (e.g., administering antibiotics if patients do not have sepsis). Our simulations, as detailed below, consider various levels of *α*, allowing for comparisons across different parameter assumptions. We also include parameters in the model that characterize physician adherence to sepsis alarms and tolerances to false alarms (i.e., over-treatment).

For simplicity, our model implicitly assumes that most sepsis cost is associated with the downstream consequences of sepsis, such as organ failure, need for intensive care, and prolonged hospitalization^15^. As such, we assume the costs of broad-spectrum antibiotics and other early sepsis treatments are negligible and thus excluded from our analysis^30^.

### Simulations

We simulate a series of outcomes by implementing sepsis prediction algorithms with flexible classification thresholds. Simulation parameters and definitions are provided in Table 1.

**Table 1.**
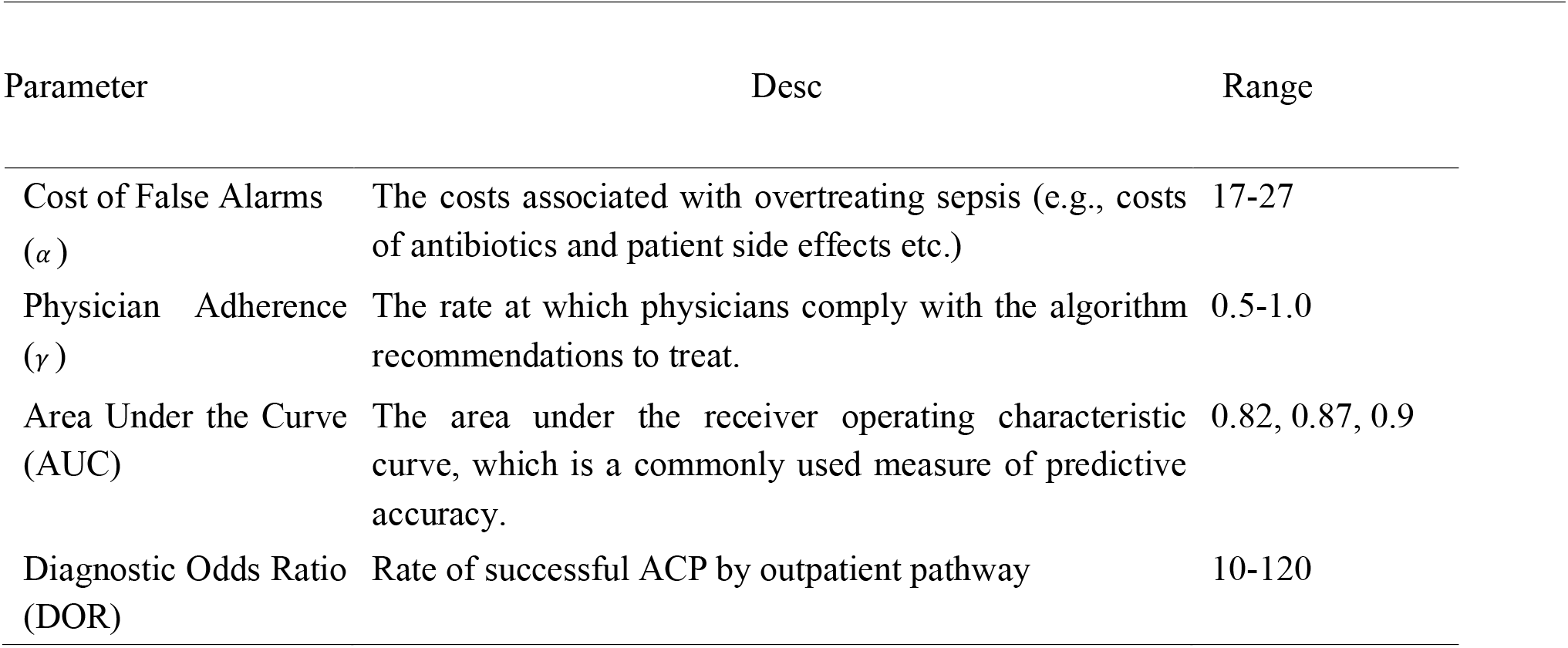
Simulation parameters

#### Model performance and false alarm tolerance

The first simulation illustrates the cost savings generated when choosing classification thresholds across diagnostic categories. The simulation presents cost savings achieved when using three different AI models^5^ with various levels of predictive performance (i.e., AUC) and a range of different tolerances to false alarms (e.g., higher tolerances mean that the costs of overtreatment are lower). This exercise illustrates the returns of allowing flexible classification thresholds across diagnostic categories for various ML algorithms and cost assumptions. We then calculate and present the diagnostic odds ratios (DOR) at each accuracy level and cost assumption given the optimized classification thresholds.

#### Physician adherence

We then reformulate the optimizer to account for physician adherence. For a given classification threshold, low adherence leads to a lower detection rate as alarms are ignored. To illustrate the effects of physician adherence on costs, we run a similar simulation to the above, but rather than considering three models of differing accuracy, we vary the adherence rate. Hence, the simulation calculates excess costs at the set of optima for different adherence parameters and costs of false alarms. Lower levels of γ indicate a lower level of physician adherence (see equation A2 in Supplementary Materials).

#### Comparison to Uniform Classification Threshold Chosen by Optimizer

We underscore the gains from optimizing classification thresholds by department. To this end, we do the same set of simulations when allowing only one classification threshold across departments. We then calculate the excess costs for different false alarm costs and accuracy levels at the optimal threshold. We also calculate the DORs at these optima.

#### Comparison to Uniform Classification Threshold

We calculate the excess costs if the algorithm implementers use a uniform 80% sensitivity, representing a clinically useful target detection rate^5^. We calculate excess costs at different false alarm costs, physician adherence, and accuracy levels, and we calculate the DORs at the optima.

## RESULTS

### Calculation of Excess Costs of Sepsis

Figure 2 shows the distribution of mean excess inpatient sepsis payments by DRG. The distribution’s mean is $23,929, and its median is $8,124. Importantly, this implies that, on average, sepsis patients generate $24,000 more charges than non-septic patients within the same DRG (matched on baseline severity). Differences between payments for patients within the same DRG weight exist because Medicare reimburses extra for costlier hospital encounters. Patients with high cost-to-charge ratios receive additional payments to compensate for hospital losses, called outlier payments.^31^ Thus, if the costs of sepsis treatment, or other non-sepsis treatments, exceed a certain threshold, Medicare compensates the hospital a certain percentage of the costs above the standard Medicare payment. Hence, outlier payments drive the difference in Medicare payments within the same DRG weight. Notice that outlier payments also explain why some sepsis patients are less costly than non-septic patients: outlier payments for these non-septic patients happen to be higher for other care unrelated to sepsis. The aggregated excess costs by MDC category are presented in Table A1 in the Supplementary Materials section. This table shows that sepsis can additionally cost Medicare up to $85,000 per patient.

**Figure 2.**
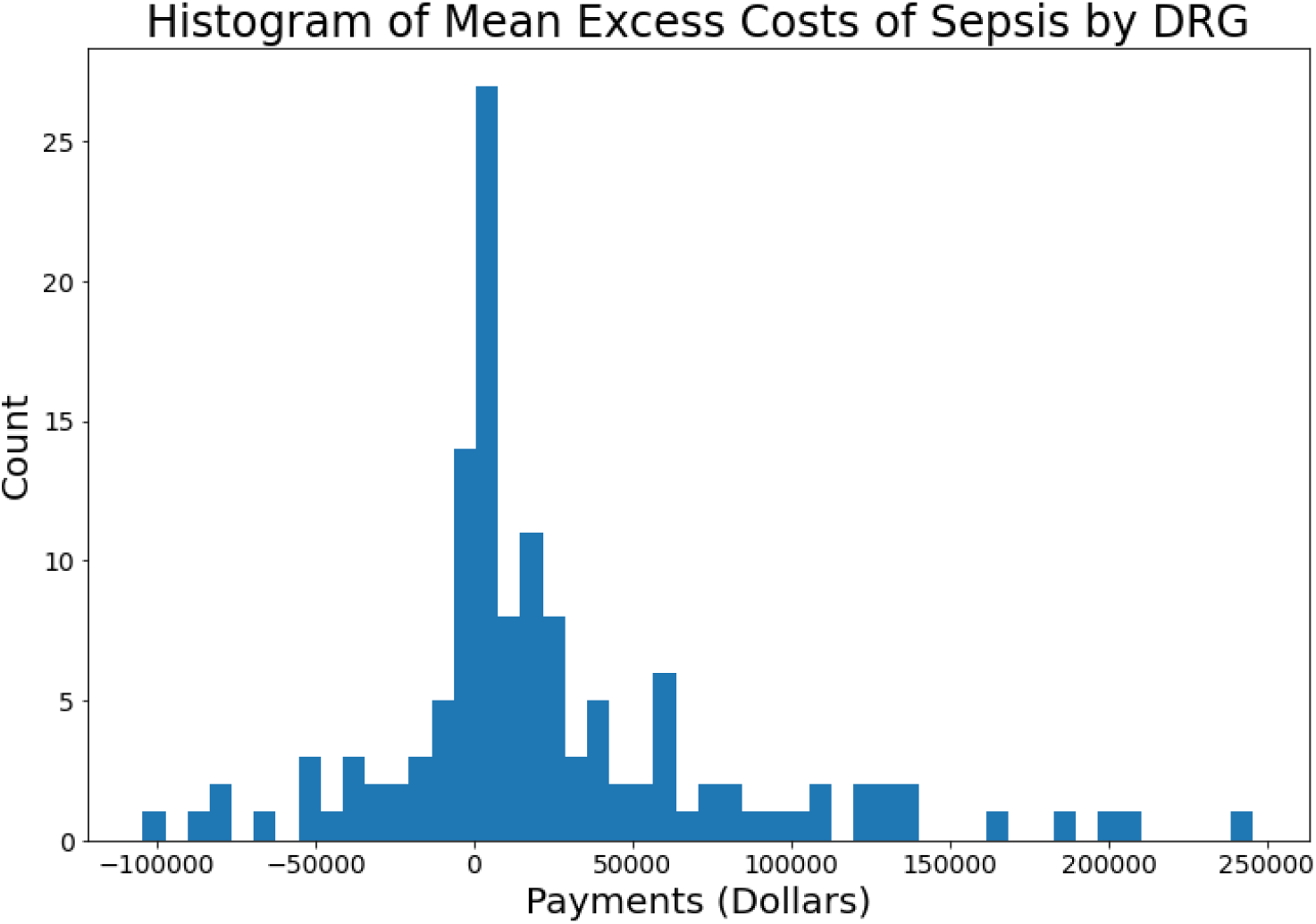
Distribution of mean excess sepsis payments over all Diagnosis-Related Groups (DRGs). This is the distribution of excess costs, as presented in Figure 3, but limited to the UCSDH cohort.

The second set of results describe the outcomes of a simulation of excess cost savings and diagnostic odds ratios achieved by ML algorithms with fine-tuned classification thresholds. We estimate that the excess cost for inpatient sepsis cases in the U.S. is $5.2 billion per year before predictive analytics implementation (see Supplementary Material section for details). Note that this estimate does not consider those patients whose primary diagnostic category is sepsis, but those who belong to non-septic DRGs who additionally have a sepsis diagnosis. The latter group incurs a total cost of roughly the same amount as the excess costs associated with our study’s patient cohort (see Figure 3). Additionally, our excess cost estimate ignores excess utilization of inpatient providers, skilled nursing facilities, and costs incurred due to the high 30-day sepsis readmission rates which is estimated to be 20% ^32,33^. Thus, our estimate likely provides a lower-bound on excess costs.

**Figure 3.**
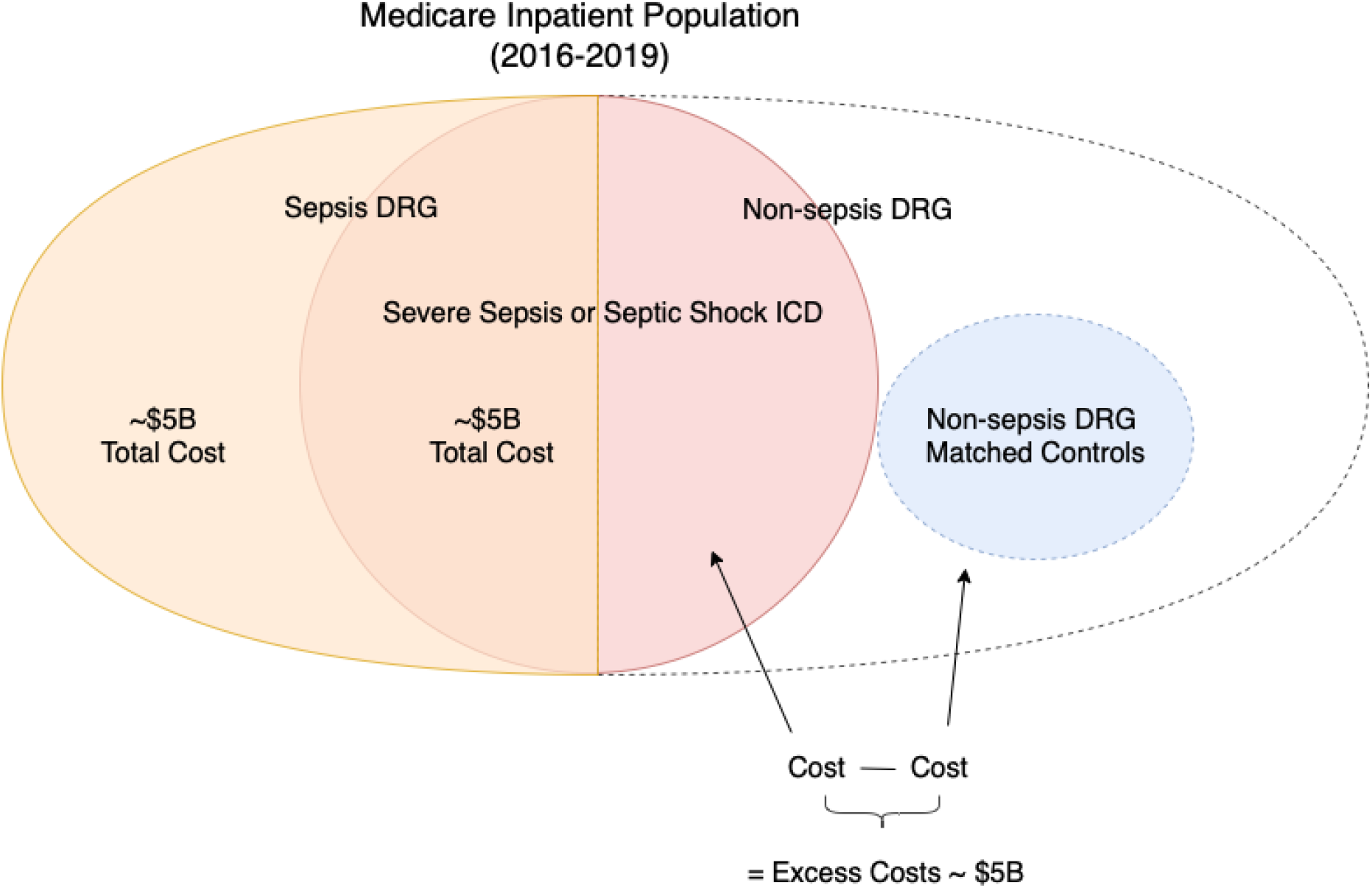
Venn diagram of Medicare Population by DRG Group and Severe Sepsis Diagnosis

Our simulation results show the savings achieved with our implementation across various assumptions. Our first set of results describes three simulation routines that differ by the degrees of freedom with which classification thresholds are chosen: (A) corresponding to 80% sensitivity, (B) a uniform across all diagnostic categories, (C) distinct and optimized for each diagnostic category. We present cost savings for each degree of freedom across various assumptions on ML accuracy and false-positive costs. Our second set of results provides the Diagnostic Odds Ratios (DOR) for degrees of freedom (A)-(C) at the optima chosen to minimize costs across the same ML accuracy and false-positive cost assumptions.

### Cost Savings

A. *Uniform Classification Threshold*. The first results detail cost savings when using a uniform recommendation of 80% sensitivity and applying it throughout the hospital at different false-positive costs and various levels of ML accuracy. Note, this implementation differs from the two others as the threshold is not optimized. Panel A, Figure 4 shows that as the cost of false positives decreases (i.e., higher α values), classification thresholds are chosen more aggressively, which leads to higher cost savings as more septic patients are diagnosed and treated. Similarly, as the predictive power of the model increases (i.e., higher AUC), savings increase. The most influential factor in cost savings is the model’s predictive power, with excess cost savings ranging from $2.3-$3.9 billion.
B. *Uniform Classification Threshold Chosen by Optimizer*. Instead of relying on a uniform recommended threshold, implementers may choose one to minimize costs throughout the hospital. The simulation of this implementation shows to what extent cost savings would differ. Panel B, Figure 4 shows that, for every AUC-α pair, cost savings are higher when the threshold is chosen. Where savings are highest, an optimized uniform threshold can save over $400 million relative to the uniform recommended level ($3.9 billion cost savings with 80% uniform and $4.3 billion cost savings with uniform chosen). Cost savings exhibit similar patterns across α and AUC values as the above model.
C. *Heterogenous Classification Thresholds Chosen by Optimizer*. Further, implementers could optimize thresholds across broad diagnostic categories (or hospital departments). Panel C, Figure 4 shows that the gains from choosing heterogenous thresholds by MDCs are highest for lower-accuracy models. This discrepancy is illustrated by the difference between cost savings in the uniform model versus the heterogeneous model, with cost savings at AUC of 0.82 as high as $3.7 billion when using heterogeneous thresholds compared to $3 billion with the uniform model. At the pair where the highest savings are achieved, heterogeneous thresholds can save over $300 million relative to uniform thresholds ($4.3 billion cost savings with uniform and $4.6 billion with heterogeneous) and almost $700 million compared to the 80% standard. Cost savings exhibit similar patterns across α and AUC values as the above models.
D. *Comparison of Cost Savings Across Degrees of Freedom*. Panel D of Figure 4 shows that heterogenous thresholds would increase cost savings by almost $700 million each year, relative to 80% uniform thresholds and by as much as $300 million each year, relative to a uniform chosen threshold. These calculations assume an AUC of 0.9 and an α of 20 across the three models. An α of 20 aligns with the maximum penalty for false alarms in the Physionet challenge^4^, and the AUC of 0.9 is close to the predictive accuracy of the latest advancement in sepsis predictive analytics by Shashikumar at al. ^5^

**Figure 4.**
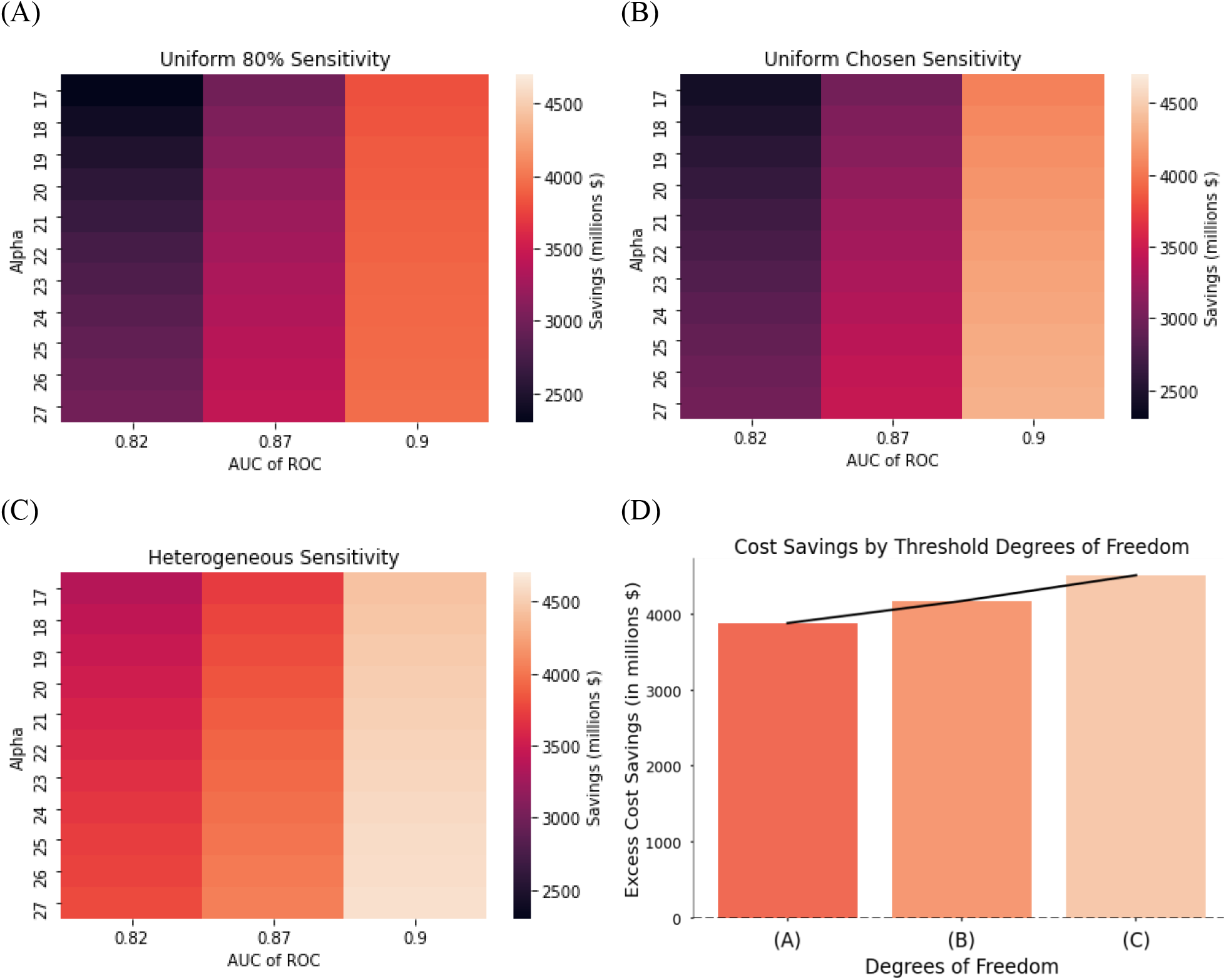
Note: The cost savings estimates in the bottom-right subfigure are for AUC = 0.9 and α = 20 across the three models. We choose α = 20 to align with the maximum penalty for false alarms in the Physionet challenge.^4^

### Diagnostic Odds Ratio

A. *Uniform Classification Threshold*. We present an objective measure of accuracy, called the Diagnostic Odds Ratio (DOR), attained at each AUC-α pair given the optimal thresholds. Panel A, Figure 5 illustrates that the highest levels of diagnostic accuracy are achieved when costs are lowest, suggesting that cost minimization can simultaneously maximize algorithmic performance. Naturally, more predictive models also lead to higher DOR values.
B. *Uniform Classification Threshold Chosen by Optimizer*. Optimizing the uniform threshold leads to higher DORs at each AUC-α pair. This improvement is prominent in most accurate models, where DOR can differ by as much as 30 between different degrees of freedom (see Panel B, Figure 5).
C. *Heterogenous Classification Thresholds Chosen by Optimizer*. Heterogenous thresholds further increase the DOR at every point relative to the previous two alternatives. In Panel C, Figure 5, we see that DOR reaches up to 116 at the highest point.
D. *Comparison of Cost Savings Across Degrees of Freedom*. Panel D of Figure 5 shows the DOR can increase by as much as 50 when switching from a uniform recommended threshold to heterogeneous thresholds, even though DOR is not directly maximized. Interestingly, minimizing excess sepsis costs also leads to higher DOR.

**Figure 5.**
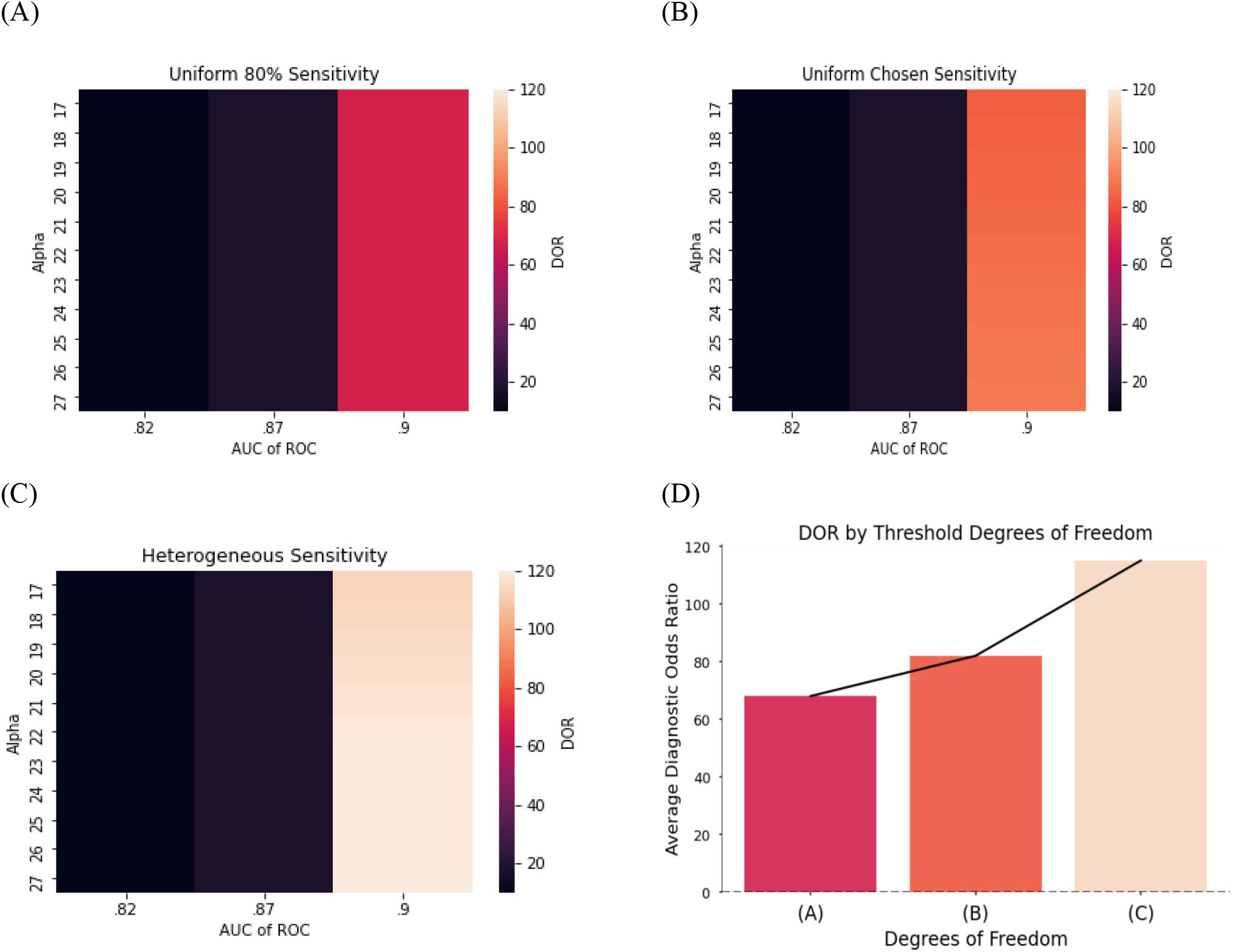
Note: The cost savings estimates in the bottom-right subfigure are for AUC = 0.9 and α = 20 across the three models. We choose α = 20 to align with the maximum penalty for false alarms in the Physionet challenge.^4^

**Figure 6.**
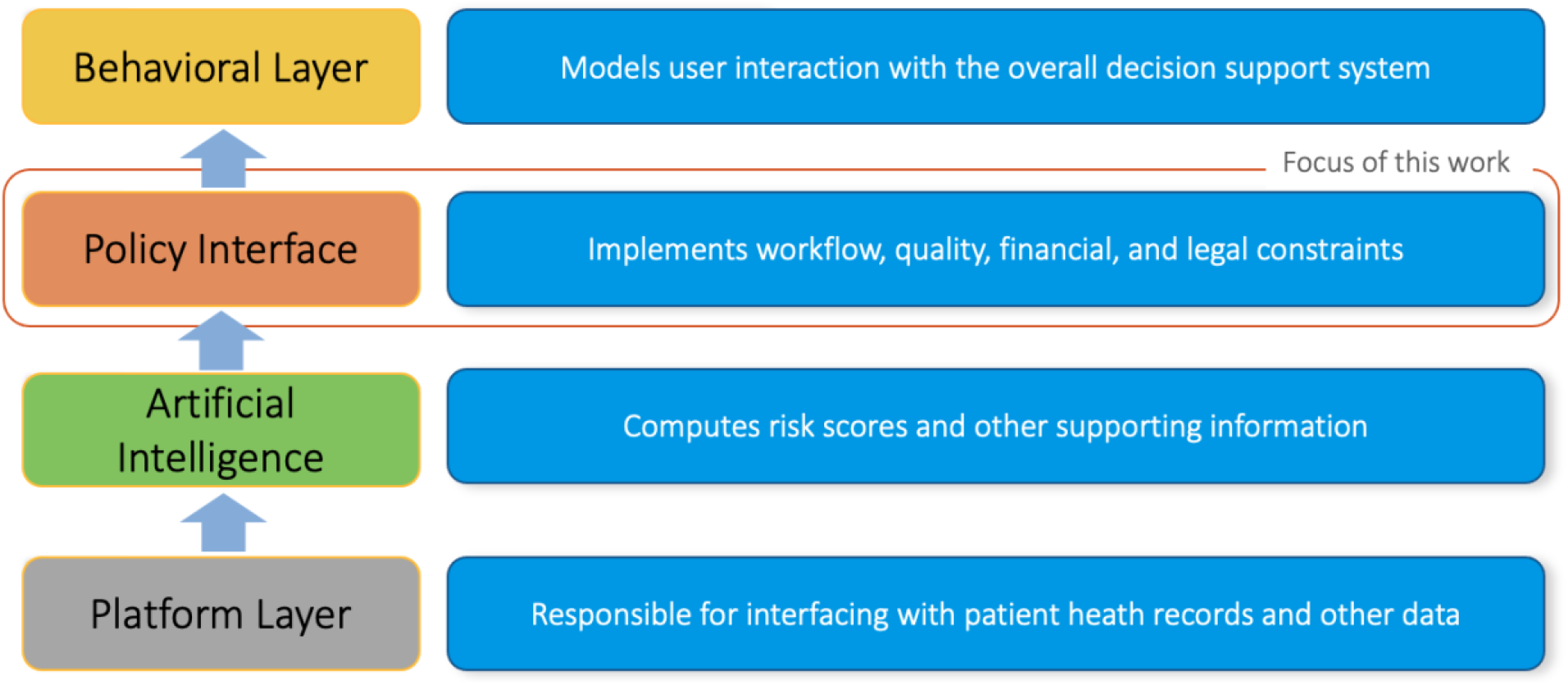
Clinical decision support (CDS) implementation layers. These layers (and the corresponding key attributes) include: 1) the Platform layer (interoperability, scalability and fault-tolerant), 2) the Artificial Intelligence layer (accuracy, generalizability, and interoperability), 3) the Policy layer (specific and applicable to local hospital workflows, optimality w.r.t. enterprise’s objectives), and 3) the Behavioral layer (usability, compliance).

### Savings when Accounting for Provider Adherence

We present results from a set of simulations that fix AUC at 0.87, but which vary the costs of false positives and physician adherence to alarm triggers. Not surprisingly, savings are highest when adherence is high (see figure A1). This result highlights the value of adequate training and quality controls to ensure that physicians and front-line workers who interact with these technologies use them appropriately. Measures to improve physician adherence to alarm triggers could increase cost savings by as much as $1 billion dollars ($3 billion at γ = .5, α = 17; $4 billion at γ = 1.0, α = 17).

## DISCUSSION

This work estimates the national excess costs of sepsis and provides a framework for implementing predictive models in clinical settings to account for these needs. Our framework chooses classification thresholds, or the points above which a patient is deemed septic, across broad diagnostic categories to minimize the costs of under- and over-treatment. We illustrate that implementing such algorithms nationwide could potentially save the Center for Medicare and Medicaid Services (CMS) over $4.6 billion each year from inpatient hospital-related costs alone. As much as 12.3% of these savings are attributable to our framework for implementation alone, relative to adhering to uniform classification thresholds. We find that diagnostic accuracy would also improve by as much as 68%.

Our work expands the frontier of research on clinical predictive models in several directions. First, we provide a methodology for calculating the excess costs of a given condition and apply that method to sepsis care. Second, to our knowledge, we are the first to provide a framework for optimizing the parameters of predictive models according to patient subpopulation. Third, our framework is the first to explicitly balance the costs of under-treatment (i.e., false negatives) and over-treatment (i.e., false positives) using a constrained optimization routine. Fourth, we allow for a flexible set of hospital-specific parameters that can be rationalized and set by the implementer. Among these, we include the possibility of imperfect adherence to triggered alarms (i.e., behavioral failures), or other factors that might influence the effectiveness of the sepsis treatments, given the alarm is followed (imperfect treatment). We also include a flexible parameter identifying the costs of false positives (i.e., over-treatment). Since this cost is a difficult value to ascertain and specific to a given hospital and/or condition, we allow the user to set this parameter at a level which they deem reasonable. We show that across various assumptions on physician adherence and over-treatment costs, our framework can dramatically increase excess cost savings.

By contrast, recent work in predictive analytics considers the cost of prediction in terms of the number of laboratory tests and their associated costs.^32^ However, these approaches overlook the more significant costs incurred from avoidable hospital expenses and insurance payouts that could be prevented by more timely and appropriate health care. Our implementation directly minimizes these costs to optimize predictive analytics.

Our approach also allows hospitals and practitioners to reap savings under current DRG-based payment models and value-based care systems.^33^ For example, under the increasingly used model of capitation payments, hospitals are allotted a payment for a fixed number of patient lives. Our implementation allows hospitals to optimize their predictive analytics within patient subgroups and provide targeted treatment depending on the needs of those subgroups. Excess cost savings from this targeted approach would be directly reaped by hospitals, incentivizing adoption of new prediction technologies.

A comparison of results across different model parameter values and inclusion criteria offers several broader insights. First, improving provider compliance to algorithmic recommendations can yield substantial cost savings. These savings are as large as those reaped when setting classification thresholds by broad diagnostic categories, highlighting the importance of dedicating time and resources to the Behavioral Layer (see Figure 5) of the clinical decision support process. By improving compliance to algorithmic recommendations and optimizing model parameters by patient subpopulation, costs can be further reduced by as much as 40%. Thus, the value proposition of new predictive models depends on how well algorithms are implemented.

Relatedly, our model could be extended to allow provider compliance rates that vary by department. These heterogenous compliance rates could, in turn, affect cost-savings outcomes. Further, one could simulate the potential savings of educational interventions that improve compliance rates within low-compliance departments.

Third, broadening the inclusion criteria of these technologies may lead to much higher excess cost savings. Our strategy, for example, only includes patients with ICD codes corresponding to severe sepsis and septic shock. By contrast, if the inclusion criteria were expanded to cover patients with any sepsis ICD code^34^ that map to the sepsis DRG codes (i.e., 870-872), the excess cost savings could double (see Supplementary Materials). Moreover, if predictive technologies were deployed beyond inpatient settings, such as in outpatient clinics, skilled nursing facilities, or via at-home wearable devices, cost savings could further increase.

Lastly, the cost of false alarms can greatly affect the potential for cost savings. If the costs of sepsis overtreatment are high relative to the costs of undertreatment (e.g., worst-case antimicrobial resistance scenarios) cost savings are limited. Identifying these costs, thus, is critical to identifying optimal classification thresholds. However, these costs could vary by hospital or department and may merit more specific calculations.

Our analysis, of course, has limitations. First, it is difficult to estimate the true excess costs of sepsis. Our estimates, which compare patients within the same DRG and with similar baseline comorbidity indices, attempt to isolate the effect of sepsis on excess costs. Our estimates, however, are an imperfect attempt at identifying the causal effect of sepsis on costs and could include other factors that increase costs apart from sepsis. Second, our analysis uses data from only one hospital. Obtaining fine-grained costs from hospitals is an arduous process, thus, we are limited by our sample size. Third, we assume that provider costs are a small fraction of procedure costs, ignoring differences in provider costs between septic and non-septic patients within MDC categories. However, septic patients often require more physician time; therefore, our excess cost estimates could be interpreted as a lower bound. Further, we do not account for the value of lives saved from improved treatment and any costs incurred after discharge, despite readmissions from sepsis being extremely common and expensive^35^. Thus, we analyzed excess costs of septic patients for whom sepsis is a non-primary diagnosis while accounting for other primary reasons for hospital admission. This allowed us to analyze avoidable costs that could be prevented by early sepsis detection during hospital care. Despite these limitations, we believe that our analysis serves a useful framework for the deployment of predictive analytics in clinical settings and underscores the potential savings when these models are deployed in a manner that directly considers costs.

## CONCLUSION

We show that fine-tuning prediction technologies to perform well under behavioral and cost constraints can improve patient outcomes while reducing health care spending. We estimate that Medicare could save over $4.6 billion each year from inpatient hospital-related costs alone, and that diagnostic accuracy would improve by as much as 68% through use of a ML-algorithm to predict sepsis. Our results suggest that the value proposition of new prediction technologies can be improved through fine-tuning within a clinical setting. Prospective studies are needed to validate these findings.

## Data Availability

The financial data underlying this article cannot be shared publicly due to the proprietary nature of the data. Center for Medicare and Medicate Provider Utilization and Payment Data (Inpatient) used for extrapolation of costs and case-mix adjustments can be found at https://www.cms.gov/Research-Statistics-Data-and-Systems/Statistics-Trends-and-Reports/Medicare-Provider-Charge-Data/Inpatient and https://www.cms.gov/files/zip/fy-2021-ipps-fr-case-mix-index-file.zip

## AUTHOR CONTRIBUTIONS

PR and SN conceived the project and the initial analysis plan. AB and SPS helped with data collection. GW and C.A.L provided domain expertise on the healthcare delivery factors. PR wrote the code and set up the simulation analysis. All authors contributed to the interpretation of the findings and contributed to the final preparation of the manuscript.

## SUPPLEMENTARY MATERIAL

Supplementary material is available at *Journal of the American Medical Informatics Association* online.

## ACKNOWLEDGMENTS

We would like to thank Julie King, director of decision support at UC San Diego Health, for providing operational context and feedback on analyses methods used in this study.

## SUPPLEMENTARY MATERIAL

### 1. Cost Minimization Routine Across Diagnostic Categories

We choose the sensitivity of the algorithm in each MDC code *i* to minimize sepsis payments subject to boundary constraints. The primary constraint is the tradeoff between sensitivity and specificity: higher sensitivity leads to lower specificity. The sharpness of that tradeoff is determined by the ROC curve. The formulation of the optimization routine is as follows:

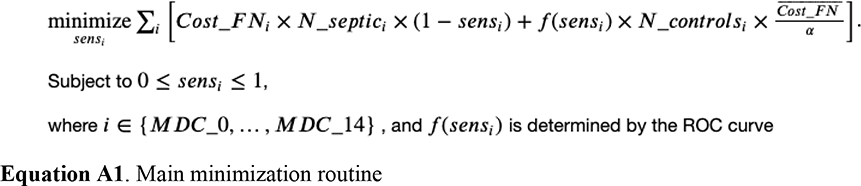

Here, *sens*_*i*_ is sensitivity and *spec*_*i*_ is specificity. *Cost_FN*_*i*_ is given by the cost of false negatives in the given MDC code *i*. This was gathered earlier by calculating the difference between CMS payments for patients with sepsis within a given DRG weight (DRG and quarter) and patients without sepsis within that same DRG weight that were matched on baseline CCI (severity). *N_septic*_*i*_ is the number of septic patients in the given MDC code and (*1 – sens*_*i*_) is the miss rate. Thus, the left-hand side of the objective function can be thought of as the total additional payments made by CMS for septic patients. *f(sens*_*i*_) is the false positive rate, which is calculated using a fitted function *f()* to the predictive model’s ROC curve. The estimated functional form *f()* thus provides a mapping from the chosen sensitivity to specificity. *N_controls*_*i*_ represents the total number of patients in the MDC code/department that do not have sepsis. The parameter *Cost_FN* bar is the average cost of false negatives across all MDC codes. The parameter *α* is a variable that maps the cost of false positives to the cost of false negatives as there may be a cost of giving someone antibiotics if they do not have sepsis (e.g., adverse side effects). Thus, the right-hand side of the objective function can be thought of as the additional cost from false positives.

### 2. Cost Minimization Routine with Imperfect Adherence

To model the cost effects of imperfect physician adherence to alarm triggers, we reformulate our model to allow a portion of true positives to be ignored (behavioral failure). Alternatively, our model extension could account for a treatment failing to prevent sepsis even if the alarm is followed (treatment failure). To this end, we add a *γ* exponent to the miss rate to scale up the miss rate for a given choice of sensitivity:

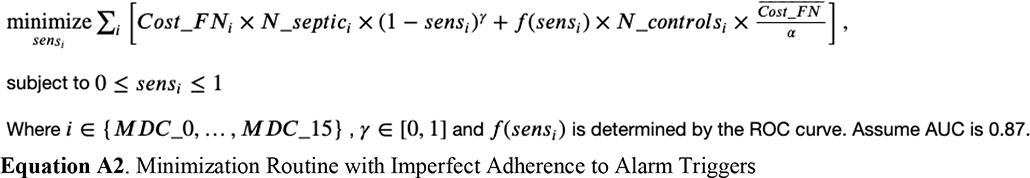

For this model, we use the model that has an AUC of 0.87. Note that the γ exponent is between 0 and 1 and the miss rate is between 0 and 1. Thus, as γ decreases, the miss rate increases.

**Figure A1.**
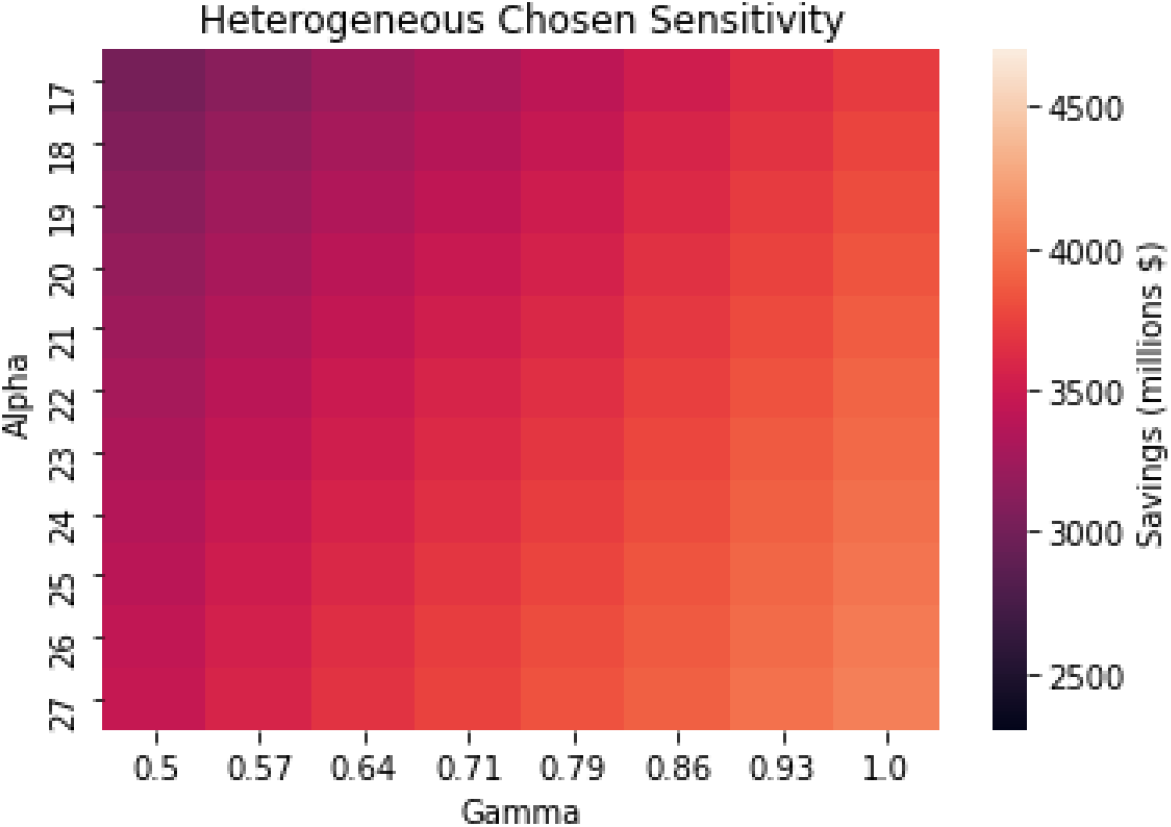
Cost Savings Across α Values and Physician Adherence Levels.

### 3. Cost Minimization Routine with Uniform Sensitivity Across All MDCs

Now we compare having the flexibility to determine sensitivity/specificity of the algorithm across departments (MDC codes) to simply choosing one uniform sensitivity/specificity pair across all MDCs.

The formulation for this optimization routine is as follows:

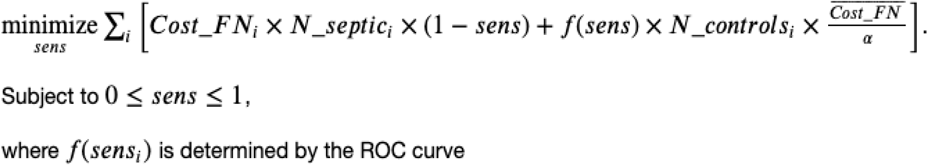

Notice that, instead of choosing a set of sensitivity values across diagnostic categories, we choose only one sensitivity value across the entire hospital.

### 4. Cost Minimization Routine With 80% Uniform Sensitivity Across all MDCs

For this simulation we simply impose a uniform 80% sensitivity across all diagnostic categories consistent with uniform, recommended levels. In this context, no algorithmic optimization for a sensitivity value is used. Costs are calculated by summing up the costs across diagnostic categories with the uniform 80% sensitivity value and the corresponding specificity value determined by the ROC curve.

### 5. ROC Curve Smoothing for AUC Constraint

To fit a function to the ROC curve, to bound the accuracy of the model, we first invert the ROC curve data points to get a mapping between *y = f(sens)*, where y is the false-positive rate, which we insert directly in the objective function. We then transform sensitivity and specificity pairs from the actual ROC curve into logit space using the following transformation

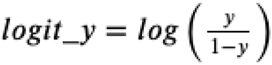

While in logit space we then fitted a model that mapped sensitivity (true-positive rate) to the false positive rate using the following regression model:

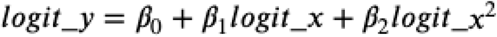

Once we obtain the fitted coefficients, we mapped each value of sensitivity to the false-positive rate by first transforming sensitivity into logit space, generating the logit false positive rate, then returning the logit false-positive rate back into the normal false-positive rate through the inverse logit function.

Using interpolation methods to estimate the ROC curve generated functions that were not smooth and that often lead the optimizer to settle on unstable solutions. With our method, we guarantee a smooth, monotonic function for every value of sensitivity.

Below we plot the original data for the ROC curve vs. the fitted data:

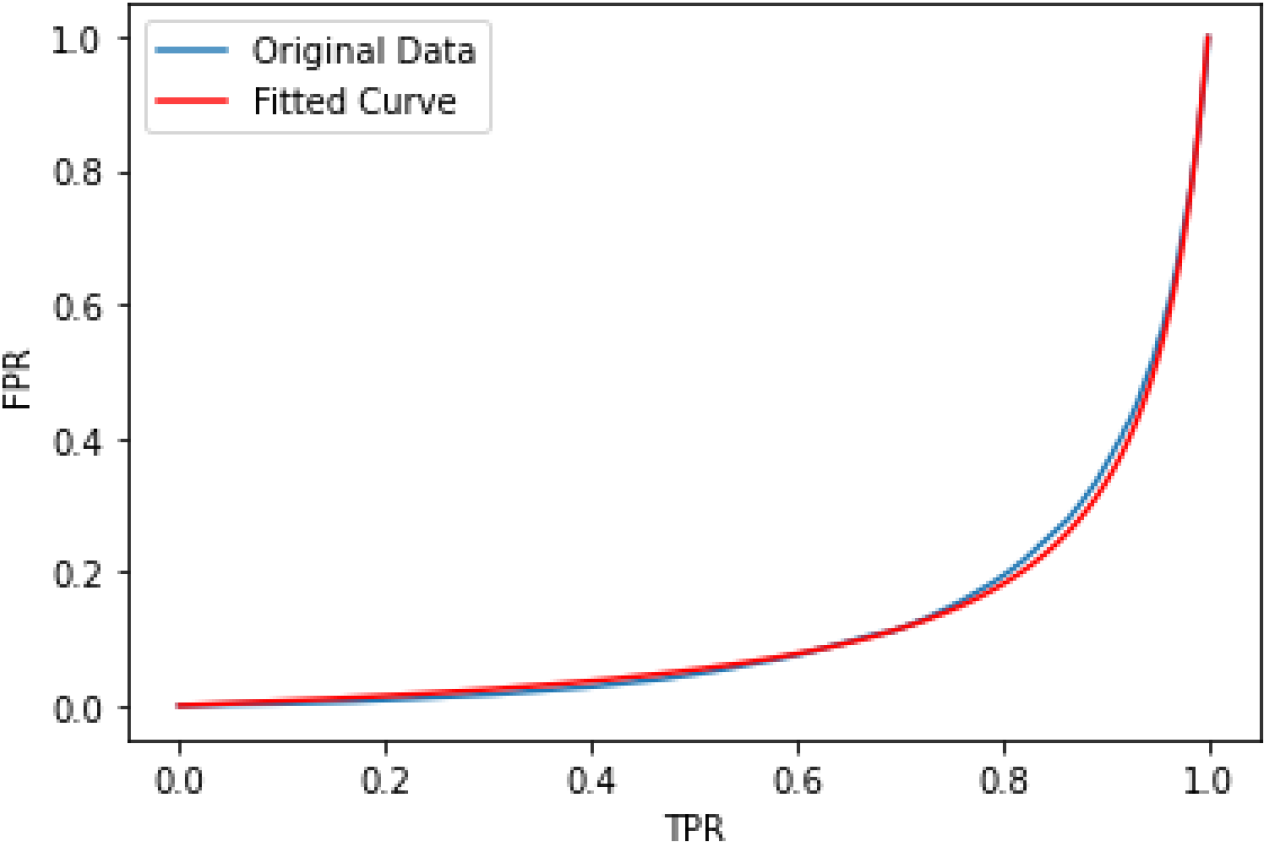

### 6. Scaling Excess Costs at UCSDH To National Level

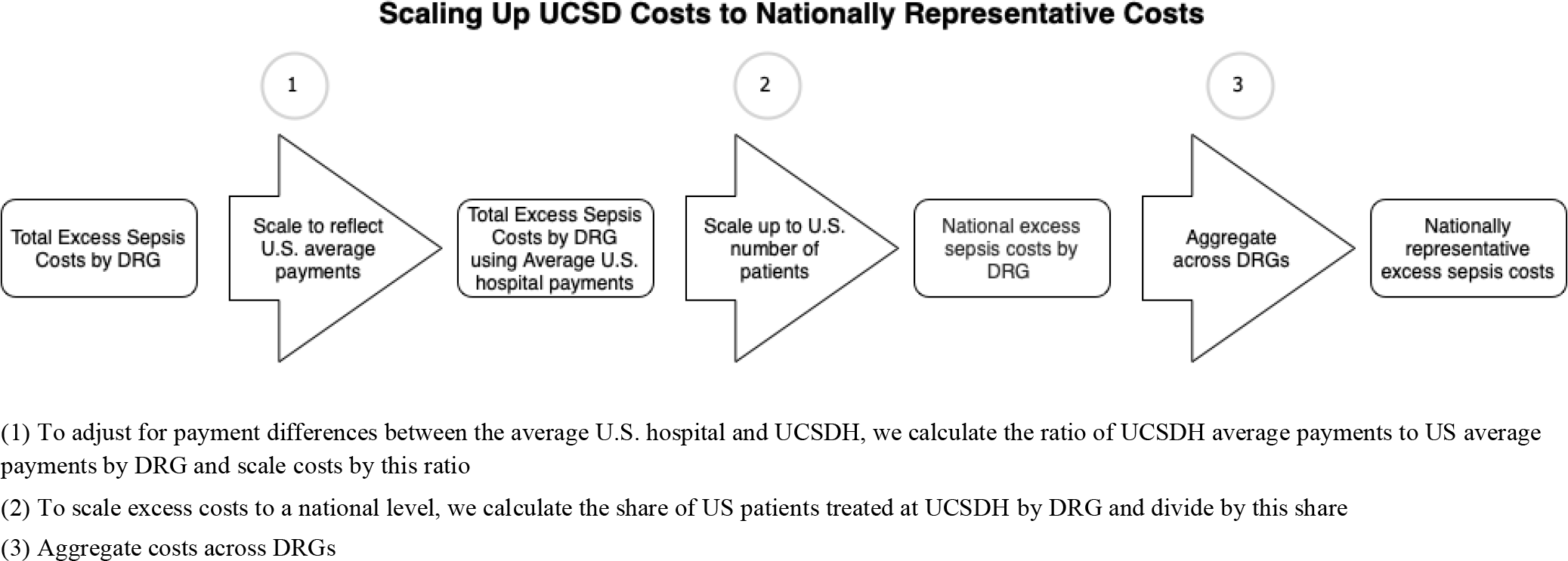

Scholars have valued the total U.S. annual costs of sepsis among Medicare Beneficiaries at $41.8B^19^. This number is derived estimating the costs from patients within the sepsis DRGs 870-872. There are two potential shortcomings of this estimate. First, it does not include sepsis diagnoses that occur in other DRG codes. By only analyzing these patients, some costs could be missed. Second, these costs are not net of the counterfactual level of care these patients would have received without sepsis. It is plausible that even if these patients did not have sepsis as a primary reason for admission, that these patients would have still been admitted to the hospital for other reasons in the near term (I.e., since these patients have more underlying conditions). Measuring the counterfactual level of costs, a patient would have incurred is essential for determining the actual cost of sepsis. That is, if we could prevent sepsis from occurring, what savings could society reap?

By analyzing patients outside the sepsis DRGs, we can do comparisons between individuals with similar underlying conditions and primary reasons for admission, but where sepsis is the key differentiator. Thus, our estimates provide a reasonable estimate of the excess costs of sepsis and can be used to scale the sepsis estimate costs in sepsis DRGs.

To scale our hospital-level estimates of excess costs to the national level, we aggregate all the excess costs of sepsis at the DRG level. Then, we calculate the ratio of UCSDH average payments to US average payments by DRG. This number measures the differences in payments for each DRG that UCSDH receives (e.g., teaching hospital add-on payments). We scale the excess cost totals in each DRG by these ratios to achieve a more nationally representative excess cost measure at the hospital level. Next, to scale these costs up nationally, we calculate the share of national patients that UCSDH treats in each DRG. We then divide the payment-adjusted DRG-level excess costs by these shares to achieve a national DRG excess cost. These costs, however, could be biased by UCSDH’s case mix. Thus, we scale these costs by Medicare’s case-mix index to normalize these costs to represent the national case mix.

The results of this estimation suggest that the excess costs of sepsis in non-sepsis DRGs is $5.2 billion annually. Note that, in our data, there is a similar number of patients within non-sepsis DRGs than within sepsis DRGs. Thus, total sepsis cost estimates may be overestimated by a factor of 10. There are, however, other costs of sepsis not measured in our data. Patients, for example, may receive antibiotic prescriptions from pharmacies, which we do not measure. Assuming we only capture half the costs of sepsis using only inpatient data, this still would amount to a 4-fold overestimation of sepsis costs. However, by including non-sepsis DRGs, the total excess costs may be somewhere close to $10 billion annually ($5 billion in non-sepsis DRGs, scaled by 2 from mismeasurement of non-inpatient costs, and $5 billion in sepsis DRGs).

Further the total number of patients in non-sepsis DRGs with sepsis ICD codes is over three times as large as the number of severe septic patients, which could double the excess cost estimates.

### 7. Validation of Scaling Exercise

To validate our scaling exercise, we focus on total costs incurred in the sepsis DRGs. As a benchmark, we use the Medicare Provider Utilization and Payment Data (Inpatient) the U.S.^29^ (“External Dataset”) and internal costs and Medicare reimbursement data from UCSDH (“Internal Dataset”). Focusing on the latter dataset, we scale the estimates of total costs for sepsis DRGs to the national level (“scaled estimates”), using the same methodology described in the section above. We then compare the total costs of sepsis from the external dataset (i.e., “the true reported values”) to the scaled estimates. We find that these costs are virtually identical. We also find that, among all sepsis patients with DRGs 870-872 (as opposed to severe septic and septic shock group included in our analysis), our scaled sepsis cost estimate using UCSDH healthcare data is the same as the scaled estimate using Medicare cost data for UCSDH. Together, these comparisons reinforce the credibility of our scaling exercise and the accuracy of our UCSDH healthcare data extract.

## MDC Table

**Table A1.** The optimal sensitivity/specificity pairs presented are evaluated at AUC = 0.90 and α = 20 for the heterogeneous chosen model. N/A pairs signify MDC categories for which there were negative excess costs associated with sepsis. This negative excess cost may reflect early death or transfers, or other factors that may correspond to lower CMS payments. We omit these categories from our analysis.

| MDC Description | Share Septic | N Septic | Excess Sepsis Costs | Optimal Sens./Spec. |
| --- | --- | --- | --- | --- |
| Pre-MDC | 0.07 | 88 | \$85739.5 | 0.98/0.75 |
| Diseases & Disorders of the Nervous System | 0.04 | 23 | \$2451.91 | 0.67/0.97 |
| Diseases & Disorders of the Ear, Nose, Mouth & Throat | 0.13 | <5 | \$64266.0 | 0.98/0.69 |
| Diseases & Disorders of the Respiratory System | 0.04 | 38 | \$4883.84 | 0.75/0.96 |
| Diseases & Disorders of the Circulatory System | 0.02 | 104 | \$29151.76 | 0.89/0.91 |
| Diseases & Disorders of the Digestive System | 0.04 | 47 | \$32531.72 | 0.93/0.87 |
| Diseases & Disorders of the Hepatobiliary System & Pancreas | 0.04 | 28 | \$18242.07 | 0.9/0.9 |
| Diseases & Disorders of the Musculoskeletal System & Connective Tissue | 0.1 | 23 | \$7827.39 | 0.91/0.89 |
| Diseases & Disorders of the Skin, Subcutaneous Tissue & Breast | 0.2 | <5 | \$7265.0 | 0.94/0.85 |
| Endocrine, Nutritional & Metabolic Diseases & Disorders | 0.04 | <5 | \$25850.5 | 0.92/0.88 |
| Diseases & Disorders of the Kidney & Urinary Tract | 0.02 | 70 | \$-23270.1 | N/A |
| Diseases & Disorders of the Male Reproductive System | 0.25 | <5 | \$765.5 | 0.8/0.94 |
| Diseases & Disorders of Blood, Blood Forming Organs, Immunologic Disorders | 0.1 | 6 | \$2519.5 | 0.81/0.94 |
| Myeloproliferative Diseases & Disorders, Poorly Differentiated Neoplasms | 0.03 | 157 | \$-12859.09 | N/A |
| Infectious & Parasitic Diseases, Systemic or Unspecified Sites | 0.08 | 12 | \$-48800.17 | N/A |
| Mental Diseases & Disorders | 0.33 | <5 | \$1570.0 | 0.9/0.9 |
| Injuries, Poisonings & Toxic Effects of Drugs | 0.05 | 16 | \$13975.75 | 0.9/0.9 |
| Factors Influencing Health Status & Other Contacts with Health Services | 0.1 | <5 | \$103.0 | 0.0/1.0 |
| Multiple Significant Trauma | 0.5 | <5 | \$46164.33 | 1.0/0.43 |
| Human Immunodeficiency Virus Infections | 0.06 | 44 | \$-36037.73 | N/A |

## Footnotes

1 Note that matching estimators do not guarantee conditional independence (or causality). However, data and institutional factors limit the applicability of causal methods in health care settings, leading us to use a matching estimator to approximate excess costs for the sake of simulation.

